# Improving executive, behavioural and socio-emotional competences in very preterm young adolescents through a mindfulness-based intervention: study protocol and feasibility

**DOI:** 10.1101/2021.01.19.21250087

**Authors:** Vanessa Siffredi, Maria Chiara Liverani, Mariana Magnus Smith, Djalel Eddine Meskaldji, Françoise Stuckelberger-Grobéty, Lorena G. A. Freitas, Jiske De Albuquerque, Emilie Savigny, Fanny Gimbert, Petra Susan Hüppi, Arnaud Merglen, Cristina Borradori Tolsa, Russia Hà-Vinh Leuchter

## Abstract

**Background:** (VPT) children and adolescents exhibit executive, behavioural and socio-emotional difficulties that persists into adulthood. Previous research suggests that mindfulness-based intervention (MBI) may specifically target the development of theses competences. The objective of the current study is to describe the study protocol and to evaluate the feasibility of a clinical trial on a MBI program to enhance executive, behavioural and socio-emotional competences in a cohort of VPT young adolescents.

**Methods:** 164 VPT young adolescents from 10 to 14 years old, born before 32 gestational weeks, were invited to participate in an MBI program of 8 weekly sessions in groups of up to 8 participants, lasting 1h30. Participant were enrolled in a randomised controlled trial (RCT) or in a pre-post intervention designs depending of their availability. Satisfaction and attendance measures of the MBI were collected using self-reported questionnaires and registration of attendance.

**Results:** Of the 63 participants who were enrolled in the study (38.2% of families invited to participate), 52 (82.5%) completed all assessments. Once enrolled, acceptability was high as shown by the high attendance rate in the sessions and the feedback evaluation questionnaire.

**Discussion:** To our knowledge, this is the first study to investigate the feasibility of an MBI study in VPT born young adolescents. Our findings suggest that an MBI study is feasible and show a high acceptability among participants. The use of an RCT design in our study constitutes the gold standard for testing the efficacy of such intervention in VPT young adolescents. If effective, the MBI program could potentially be a valuable tool for improving executive, behaviour and socio-emotional competences in the vulnerable VPT population.

**Trial registration:** ClinicalTrials, NCT04638101. Registered 19 November 2020 - Retrospectively registered, https://clinicaltrials.gov/show/NCT04638101.

## BACKGROUND

Every year, an estimated 15 million babies (more than 1 in 10 babies) worldwide are born prematurely, i.e. before 37 completed weeks of gestation [1]. As a result of medical advances in neonatal care over recent decades, survival rate of preterm infants has increased significantly. Nevertheless, the adverse neurodevelopmental consequences of surviving children remain a major concern worldwide. Extensive evidence shows that very preterm (VPT) children and adolescents, i.e., born before 32 gestational weeks, are at high risk for cognitive and social-emotional impairments that can persist up until adulthood [2]. Consequently, the focus of research in VPT children has shifted from increasing survival rates and characterising neurodevelopmental outcomes to improving outcomes and enhancing the quality of life for these at-risk infants [3].

A cognitive ability known as executive functioning has been evidenced to be particularly at risk of impairment in children and adolescents born prematurely. Executive functioning is an umbrella term that incorporates a collection of inter-related high-level cognitive skills for purposeful, goal-directed behaviour [4]. Executive functioning is crucial in daily life activities and closely linked to academic abilities. The model of Anderson (2002) [5] [5] conceptualises executive functions as four distinct domains: (i) attentional control; (ii) information processing; (iii) cognitive flexibility; and (iv) goal setting. Each of these domains have been found to be altered in VPT at different stages of childhood and adolescence. In regard to attentional control, deficits in selective attention, inhibition and self-regulation are widely observed in this population [2]. Difficulties related to information processing include lower speed of processing and reduced verbal fluency in VPT compared to their term born peers [2]. Reduction in cognitive flexibility can be observed with difficulties in shifting ability and working memory [2, 6]. Finally, goal setting difficulties seem to be less frequent but are still observed in VPT, especially for planning skills [7]. From a neurobiological view, the development of executive functions has been related to the maturation of the frontal lobes, especially the prefrontal cortices, regions particularly vulnerable to premature birth [8].

Other high-level cognitive skills found to be at risk of impairment in children and adolescents born prematurely are behavioural and socio-emotional competences. Socio-emotional competences are a set of effective and functional resources used to adapt to the environment, and especially the social environment. There is a relative consensus that socio-emotional competences refer to a set of skills related to the ability to interact with others, communicate effectively and to how individuals identify, express, understand, use and regulate their emotions and those of others [9]. The improvement of socio-emotional abilities during childhood and adolescence is an important aspect of cognitive development and has important implications on social behaviour and academic performances [10]. Children and adolescents born VPT show difficulties in emotional regulation capacities with an increased risk of irritability and higher degree of negative reactivity [11]. Reduced emotional expression, emotional facial recognition and social perceptual skills have also been reported in VPT children and adolescents [12]. Neuroimaging studies show specific alterations of the “social brain” in VPT children and adolescents, including structural and functional alteration of the orbitofrontal cortex, the amygdala, the fusiform gyrus and the superior temporal sulcus [13, 14].

Taken together, these findings suggest that VPT children and adolescents may benefit from interventions designed to enhance executive, behavioural and socio-emotional competences. An emergent body of evidence suggests that mindfulness-based practices are a way to support the development of both these high-level cognitive abilities during childhood and adolescence [15].

Mindfulness may be defined as intentionally paying attention to the present moment, with openness to what emerges and with a non-judgmental attitude [16]. In children and adolescents, mindfulness-based interventions (MBI) have been associated with enhancement of all subdomains of executive abilities, including attentional control, information processing, cognitive flexibility, and goal setting [17]. In addition, MBI have been associated with improved behavioural and socio-emotional abilities, increased emotion regulation via reduction in stress, anxiety and socio-behavioural problems, as well as greater empathy [18]. From a neurobiological view, MBIs have been mostly associated with modification in the structure and function of frontal regions involved in executive as well as socio-emotional processes. Adult studies have shown that frontal regions are particularly sensitive to MBI with an increase in cortical thickness in subjects who benefited from this type of intervention, as well as in expert meditators compared to non-meditators [19]. Changes in white matter properties of frontal regions have also been found in expert meditators with an increase in fractional anisotropy [19]. Functionally, there is strong evidence for changes in frontal activity during task and rest, following MBI training or in expert meditators (see Marchand (2014) for a review).

In the development of executive, behavioural and socio-emotion competencies, adolescence represents a key developmental window, marked by significant maturation of neurobiological processes that underlie these skills [20]. Several studies have shown dramatic improvements in executive functioning during adolescence [20], as well as in socio-emotional competences with an increase in affective modulation and in the ability to read emotional and social cues [21]. Nevertheless, adolescence is also a period of increased risk-taking behaviour, as well as a period of onset for emotional problems such as depression, violent delinquency and substance abuse [22].

Despite the promising role of MBI for executive, behavioural and socio-emotional competencies in children and adolescents, to our knowledge, the effectiveness of an MBI has not been assessed in a preterm population so far. The aims of the present study are to describe the study protocol and to explore the feasibility of a clinical trial on an 8-week MBI in VPT young adolescents and describe the acceptability of mindfulness practice.

From a long-term perspective, this research study aim to assess potential benefits of an MBI in VPT young adolescents on executive and socio-emotional outcomes as well as on associated brain structure and functions. A population of young adolescents from 10 to 14 years of age was targeted as representing a crucial developmental phase where they might benefit from MBI in preventing the significant mid-late adolescence emergence of common mental health disorders.

## METHODS

### Participants

VPT young adolescents from 10 to 14-year-old, born before 32 gestational weeks (between 01.01.2003 and 31.12.2008) in the Neonatal Unit at the Geneva University Hospital, Switzerland, and followed up at the Division of Child Development and Growth at the Geneva University Hospital, were invited to participate in the “Mindful preterm teens” study. Of note, specialised neurodevelopmental follow-up is part of the care offered to VPT infants and their families at the time of discharge from hospital. This follow-up offers multidisciplinary consultations (with developmental paediatricians, psychologists and physiotherapists) at the key ages of 6, 12, 18-24 months, 3 years, and 5 years. Thus, the participants in our cohort were last seen at our clinic at the age of 5 years. VPT young adolescents with severe sensory or physical disabilities (cerebral palsy, blindness, hearing loss), with an intelligence quotient below 70 and who were not French speaking were excluded. MBI training sessions were offered between January 2017 and May 2019.

### Ethics approval and consent to participate

This study was approved by the Swiss Ethics Committees on research involving humans (ID: 2015-00175). Written informed consent was obtained from a parent or guardian as well as from the participant. The Magnetic Resonance Imaging (MRI) component to the study was optional and participants could still participate in the trial if the young adolescent or his/her family did not wish to have an MRI or if participants had a contra-indication to participate in the MRI session, such as dental braces.

### Neonatal variables: data collection and definition

Neonatal characteristics were routinely recorded with a secure interface protecting confidentiality. Gestational age was based on the best estimation from early ultrasound or last menstrual period. Neonatal bronchopulmonary dysplasia (BPD) and major brain injury, such as intraventricular haemorrhage (IVH) grade 3 or 4 and/or cystic periventricular leukomalacia (cPVL), were defined as previously published [23].

### Recruitment

The recruitment phase of the study was conducted between September 2016 and January 2019.

Families of eligible participants first received an information letter about the MBI program and were invited to attend an information session given by one of the paediatricians involved in the study. Parents and their young adolescents were provided with detailed informatio about the study, including the proposed MBI, as well as about the MRI acquisition. For families who expressed their willingness to take part in the trial, a brief eligibility screening was conducted and they were recontacted by phone within two weeks to organise their participation.

Families who did not attend the information session were also invited to participate in the
study. Families who did not participate in the information session were contacted by phone by the research team. Verbal explanation and information about the trial were provided. For families who expressed their interest in the study, a brief eligibility screening was conducted and further written information was sent via mail or email. Families who confirmed their willingness to take part in the trial were then recontacted to organise their participation. Families who indicated that they did not want to participate in the study were no longer contacted.

Participants were either enrolled in a randomised controlled trial (RCT) design or in a pre-post intervention (PPI) design, depending on their availability, see Figure 1. To be enrolled in the RCT design, young adolescents had to be available for the intervention over a period of 6 months, without knowing whether they would be assigned to the intervention group (IG) or the waiting group (WG). If enrolment in the RCT design was not possible due to lack of time or organisational reasons, families were included in the PPI design.

**Figure 1.**
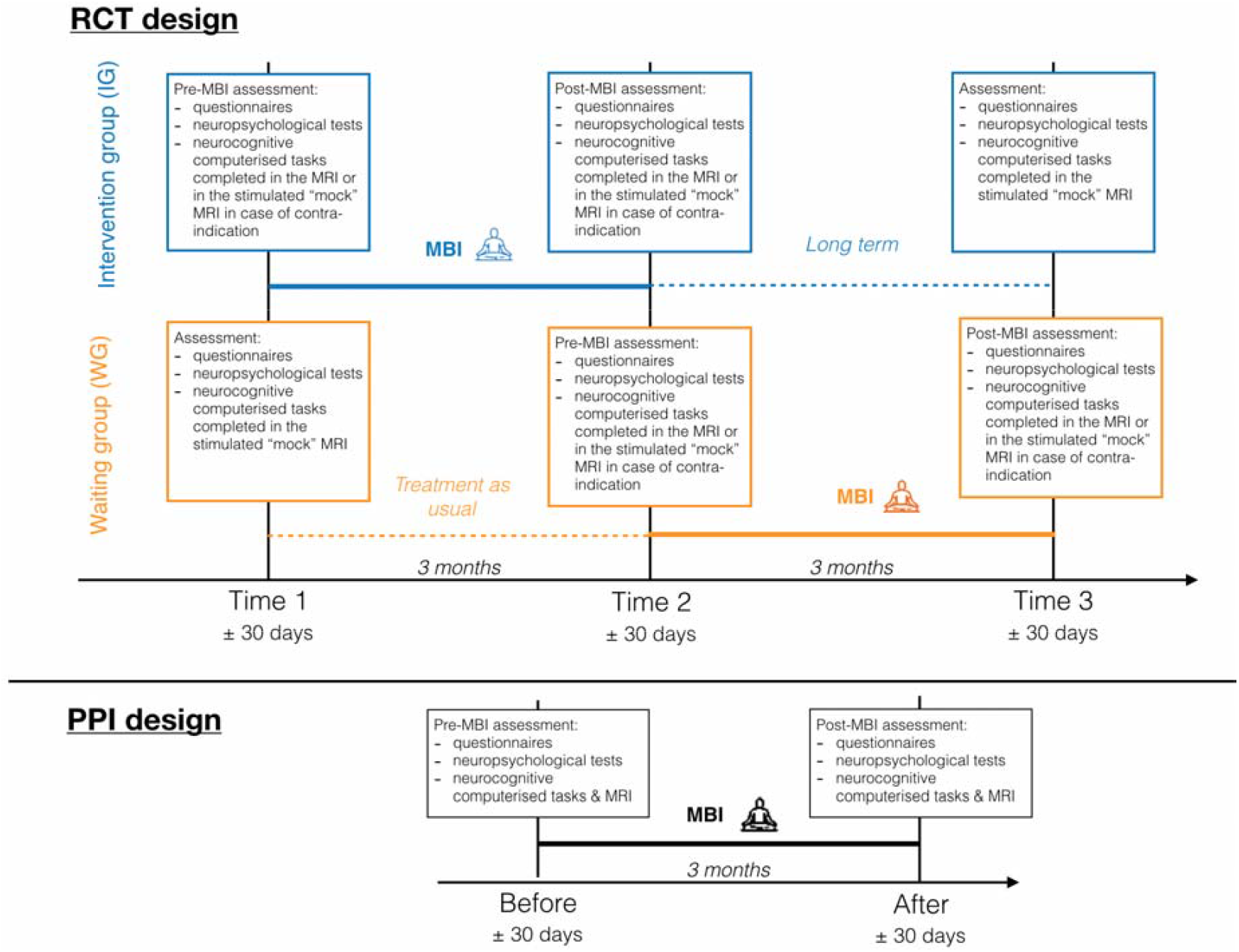
Illustration of the Randomised Controlled Trial (RCT) and Pre-Post Intervention (PPI) designs of the study. Young adolescents enrolled in the RCT design were randomised in two groups: the intervention group (IG) in blue, and the waiting group (WG) in orange. Young adolescents enrolled in the PPI design completed two assessments, one before and one after MBI.

For those enrolled in the RCT design, families were randomised either to the IG or the WG. An independent biostatistician generated a random number table. Families were allocated to the next available sequential study number which corresponded to an opaque sealed envelope which contained the randomisation allocation to the IG or WG. The project coordinators or research assistants opened the envelope to obtain the group allocation after enrolment and before the first appointment. To facilitate the participation of families with twins, the randomisation was completed for a single family, so that siblings would be consequently allocated in the same group.

Based on the initial number of VPT young adolescents from 10 to 14-year-old eligible and invited to participate, we anticipate that about 60 will participate to the present study. This estimated sample size was in accordance with our power calculation so that a clinically significant difference can be found on the effect of the MBI. Inclusion of 60 children (30 per arm in the RCT part of the study) would allow to detect an effect of MBI with a significance level of 0.05 and a power of 0.86 [18].

### Intervention

The proposed MBI was designed by the authors, adapting well-known MBI programs including Mindfulness-Based Stress Reduction (MBSR; [16]) and Mindfulness-Based Cognitive Therapy (MBCT; [24]) to adolescents’ needs and language. The length of sessions and practices were also adapted to this population. The program consisted of 8 weekly sessions in groups of up to 8 participants, lasting 1h30. Two MBI groups were offered per week (Wednesdays and Fridays) and participants had the possibility to choose the most convenient day for them. Two instructors were present for each group throughout the intervention.

For each session one theme was addressed, such as attention and the stabilisation of the focus of attention, bodily sensations, breath, emotions, thoughts, compassion, stress, stress reactivity and coping strategies. Different formal meditation practices were introduced, such as sitting mediation with different objects of attention, body scan, walking meditation and mindful movement. Participants had the opportunity to share experiences with their peers and the instructors during each session, allowing the recognition of individual patterns of behaviour, and at the same time, the appreciation of the shared experiences as a community. They were also invited to practice between 5 to 20 minutes per day at home, using guided mediation recorded by the instructors. The details about each session can be visualised in Table 1.

**Table 1.** Theme addressed during each session of the MBI intervention

| Session | SESSION THEME<br>Session Intention<br>Session Mindfulness Attitude | Agenda | Home practice |
| --- | --- | --- | --- |
| **1** | **ATTENTION and AUTOPILOT** |  |  |
|  | Introduction to attention and autopilot mode<br>Beginners Mind | Group and instructor introduction<br>Mindfulness definition: *To pay attention to what happens in the present moment, with curiosity and non-judgmentally.*<br>Dialogue about attention awareness and focus of attention<br>The 6 channels: 5 senses and thoughts<br>Practice: Eating a raisin as an explorer<br>Practice: Grounding meditation<br>Closure practice | Chart: Attention - where is my attention now?<br>Practice: Mindful eating – a mindful bite once a day<br>Practice: Grounding meditation |
| **2** | **DISCOVERING THE BODY LANGUAGE** |  |  |
|  | Discovering bodily sensations<br>Acceptance | Opening practice: Grounding meditation<br>Dialogue about home practice<br>Dialogue about sensations and sensation awareness<br>Practice: Lying down Body scan, with a component of contraction and relaxation at the beginning<br>Practice: Seated body scan<br>Closure practice | Chart: Cool moment of the day<br>Practice: Doing mindfully something habitually done on autopilot<br>Practice: Grounding meditation<br>Practice: Body scan |
| **3** | **ATTENTION STABILIZATION** |  |  |
|  | Discovering the breath<br>Non-striving | Opening practice: Grounding meditation<br>Dialogue about home practice<br>Dialogue about the breath and it's use as a possible anchor<br>Practice: The 3 minutes' break<br>Practice: Stop and breath<br>Practice: Yoga posture<br>Closure practice | Chart: Bad moment of the day<br>Practice: Body scan<br>Practice: 3 minutes' break<br>Practice: Stop and breath<br>Practice: Yoga posture |
| **4** | **RECOGNIZING EMOTIONS** |  |  |
|  | Recognizing emotions from bodily sensations | Opening practice: Grounding meditation<br>Dialogue about home practice<br>Practice: Seated body scan, including emotion<br>Dialogue about emotions: recognizing emotions from bodily sensation<br>Practice: Sitting meditation: after remembering a difficult moment, recognize the emotion(s) and it'-s manifestations<br>Drawing and naming the identified emotion<br>Closure practice: The 3 minutes' break | Chart: Emotions - recognizing links between sensations, thoughts and behavior<br>Practice: Yoga posture<br>Practice: The 3 minutes' break<br>Practice: Sitting meditation (emotions) |
| **5** | **RECOGNIZING THOUGHTS** |  |  |
|  | I'm much more than my thoughts<br>Non-judging | Opening practice: Grounding meditation<br>Dialogue about home practice<br>Walking down the street exercise<br>Discussion about thoughts and emotions<br>Practice: walking meditation | Chart: Cool moment of the day - noting sensations, thoughts and behavior<br>Practice: Sitting meditation (emotions)<br>Practice: Walking meditation |
|  |  | Closure practice |  |
| **6** | **AUTOMATIC REACTION OR CONSCIOUS RESPONSE?** |  |  |
|  | Exploring stressors and stress reaction<br>Letting go | Opening practice: Grounding meditation<br>Dialogue about home practice<br>Quick board game: identifying stress reaction<br>Discussion about stressors and stress reaction strategies<br>Practice: 5 minutes to deal with stress<br>Closure practice | Chart: Bad moment of the day - noting sensations, thoughts and behavior<br>Chart: Identifying qualities on others<br>Practice: 5 minutes to deal with stress<br>Practice: Walking mediation |
| **7** | **KINDNESS** |  |  |
|  | Being kind to oneself and to others<br>Gratitude and generosity | Opening practice: Grounding meditation<br>Dialogue about home practice<br>Discussion about kindness and compassion<br>Practice: finding refuge<br>Drawing our refuge<br>Mindful listening exercise<br>Dialogue about communications and social media<br>Closure practice | Letter to oneself:<br>What did I learned and I don't want to forget?<br>What did I learn about myself?<br>Which meditations do I want to keep practicing?<br>Practice: Refuge<br>Practice: Chose another meditation to practice |
| **8** | **CLOSURE AND OPENNESS** |  |  |
|  | Integrating the program<br>Trust | Opening practice: Grounding meditation<br>Dialogue about home practice<br>Practice: sitting meditation about the program<br>Satisfaction questionnaire<br>How to facilitate our own practice<br>Closing ritual |  |

### MBI satisfaction and attendance measures

VPT participants completed a satisfaction questionnaire during the last session of MBI. This included a question on how much they found the MBI program important to them that was used as a reference measure. In addition, the number of sessions attended were registered.

### Statistical analyses

Recruitment and drop-out rate are reported using descriptive analyses on the number of eligible participants, participation rates and retention rates in the RCT and the PPI design. Demographic, neonatal and participant characteristics are summarised by using mean and standard deviation for continuous variables and number and percent for categorical variables. For participants enrolled in the RCT, group differences between the IG and the WG are explored on demographic, neonatal and participant’s characteristics using two-tailed t-test for independent samples for continuous variables and chi-square tests or Fisher’s Exact test as appropriate for cross-tables in categorical variables. P values are reported, as well as q values corrected for all multiple comparisons completed in the present article using a False Discovery Rate correction (FDR, q<0.05) [25]. Satisfactory measure is reported using mean and standard deviations and attendance measure using percentage.

### Description of study protocol

#### Procedure

All assessments were completed at the Campus Biotech, Geneva (Switzerland). Participants from the RCT and from the PPI designs completed a baseline assessment. The baseline assessment included: a) parents and self-reported demographic questionnaires used to assessed socio-economic status and general characteristics of the young adolescent; and (b) assessment of intellectual functioning using the Wechsler Intelligence Scale for Children – 4th Edition (WISC-IV; [26] used to measure the General Ability Index (GAI; mean for the general population = 100, standard deviation = 15). Socioeconomic status of the parents was estimated using the Largo scale, a validated 12-point score based on maternal education and paternal occupation (range 2–12; [27] Higher socio-economic status scores reflect lower socio-economic level.

For participants enrolled in the RCT, three additional assessments were completed at three different time points: Time 1, Time 2, Time 3; see Figure 1. Outcome measures of the research study were collected at each different time point via parents and self-reported questionnaires, neuropsychological assessment, neurocognitive computerised tasks, a brain MRI and the recording of physiological responses for one of the computerised tasks (Emotion Regulation task; recoding of heart rate, skin conductance, respiration). Participants from the IG participated in MBI between Time 1 and Time 2. Participants from the WG took part in the MBI between Time 2 and Time 3. Participants wearing dental braces did not complete the MRI part of the study, as it produces signal distortion on MRI images.

For participants enrolled in the PPI design, two assessments were completed: one before MBI and one after MBI; see Figure 1. Research study outcome measures were identical to the RCT for the time points before and after MBI.

For all participants involved in the study, the pre-intervention assessment (i.e. Time 1 for the immediate IG, and Time 2 for the WG) was completed within 1 month before the first MBI session. The post-intervention assessment (i.e. Time 2 for the IG, and Time 3 for the WG) was completed within 1 month of the last MBI session. For participants enrolled in the RCT, the remaining assessment (i.e. Time 1 for the WG, and Time 3 for the IG) was completed 3 months before the pre-intervention assessment (for the WG) or 3 months after the post-intervention assessment (for the IG).

A gift card of 100 Swiss francs in a retail chain selling cultural and electronic products was offered to all participants who completed the study at their last appointment.

#### Neurobehavioural Assessment

Participants’ executive, behavioural and socio-emotional functioning was assessed using parents’ questionnaires, self-reported questionnaires, neuropsychological testing and neurocognitive computerised tasks. See Supplementary Table S1 for details of the research study neurobehavioural assessment.

#### MRI

Neuroimaging acquisition between 1 day and 1 month pre-intervention and between 1 day and 1 month post-intervention was performed using a Siemens 3T Magnetom Prisma scanner. During each MRI session, different types of data were acquired:

i. High resolution structural T1-weighted MP-RAGE (Magnetization Prepared Rapid Gradient Echo) sequence.
ii. Functional images were T2*-weighted with a multislice gradient-echo-planar imaging (EPI) sequence, including Resting-State fMRI data for which participants were asked to lie still with their eyes closed and engage into mind wandering and task-related activation paradigm (Flanker Visual Filtering Task, Orbitofrontal Reality Filtering task, Emotion Regulation task and Recognition of Emotions in Contextual Scene Task).
iii. Diffusion-weighted imaging (DWI) sequences were acquired with 1.3 mm3 isotropic voxels with four different shells.

Participants were carefully instructed about the imaging process and they all completed a simulated “mock” MRI session prior to the pre-intervention MRI scan. This preparation process was conducted by trained research staff and allowed participants to familiarise themselves with the scanner and the scanning process, eventually raising any concerns they might have had prior to the MRI scan. Furthermore, this process facilitated acquisition of good quality MRI images. In addition, framewise displacement of the functional sequences were calculated in real-time and framewise displacement graphs were projected onto a screen to the research staff. This allowed the research staff to give direct feedback of the participants’ motion during scanning session.

### Data management

Data were anonymised by assigning study IDs to each child and family taking part in the study. Data for all participants are stored in coded form. All data and records generated during this study are kept confidential in accordance with institutional policies and the Investigator and other site personnel will not use such data and records for any purpose other than conducting the study. Access to all data will be controlled by the PI. Hard copies of case report forms and source data are stored in a locked cabinet in a locked office. Electronic source data are stored on a network share drive with access controlled by the principal investigator. Quality control of the data incorporate range checks and between-variables consistency. Data checks for errors will be performed prior to analysis.

### Data monitoring

The incidence of adverse events was expected to be low in this single-site minimal risk research. Indeed, there was no harm as a consequence of participation in this study. The principal investigator was responsible for monitoring the data and safety of all participants. In addition to obtaining ethics approval and the data management procedures outlined above, the principal investigator hold bimonthly study team meetings to evaluate the safety and progress of all research procedures.

### Dissemination of results

Results from the study will be published in peer-reviewed academic journals as well on scientific meetings and social media. A final scientific report will be provided to the funder of the study. A plain summary which we will send to all participants at the end of the study.

## RESULTS

### Recruitment feasibility

One hundred and sixty-five potentially eligible young adolescents born VPT were invited to attend an information session on the “Mindfulness-based intervention study”.

Sixty-seven VPT young adolescents (40.6%) and their families attended the information session. Out of the families who participated in the information session, 17 (25.4%) refused and 1 (1.5%) failed to meet inclusion criteria. 52 (41.8%) families who did not participate in the information session refused to take part, 27 (17%) were unreachable and 4 (3%) failed to meet inclusion criteria. Of all the families contacted, the final participation acceptance rate was 38.2% (n = 63), and 31.5% (n = 52) completed all assessments at the different time points of the study. After enrolment in the MBI study, 11 participants (17%) dropped out before the start of MBI, including 3 (4.8%) who were unreachable, 5 (7.9%) who dropped out for lack of time and 3 (4.8%) for lack of interest. There was no drop-out after the start of the MBI program. The participant flow is illustrated in Figure 2.

**Figure 2.**
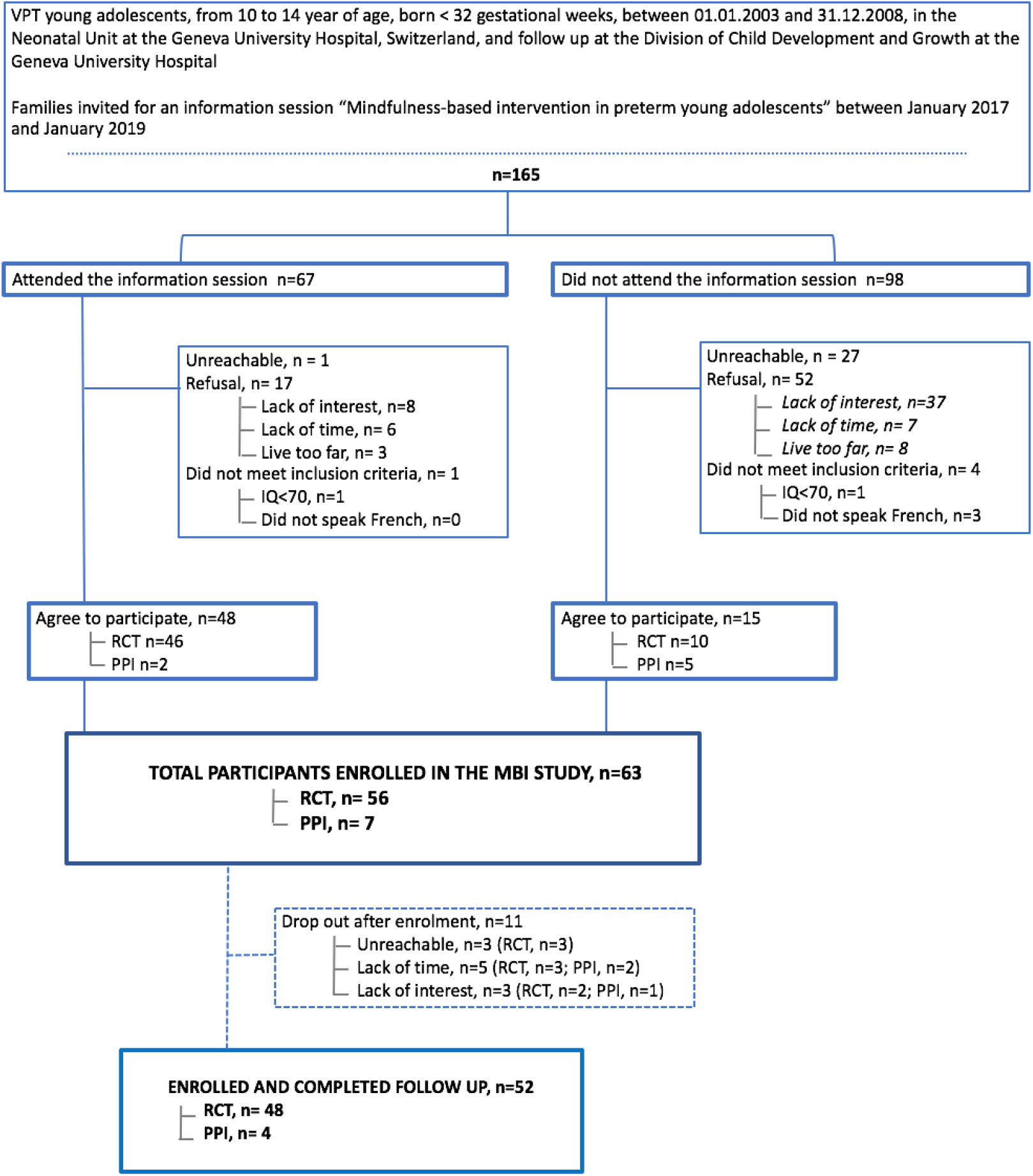
Participants’ recruitment and retention flow. Briefly, VPT young adolescents who attend in the information session about the study (n = 67) and those who did not attend the information session (n=98) were invited to participate in the current study. A total of 63 VPT young adolescents accepted to enrol in the study and 52 completed the follow-up.

Neonatal and demographic characteristics of VPT young adolescents enrolled in the MBI study (n = 63, 38.2%) and those who refused to participate (n = 102, 61.8%) are shown in Table 2. There was no significant difference between the two groups in terms of birth weight, gestational age, head circumference and presence of brain lesions linked to cystic periventricular leukomalacia. Nevertheless, there was a significant difference for multiple births, with a higher number of multiple births in the group enrolled in the MBI study compared to the group who refused to participate (q = 0.024). In regard to demographic characteristics, there was no group difference in terms of gender. Nevertheless, there was a significant group difference in parents’ socio-economic status, with significantly higher socio-economic status, i.e. lower score on the Largo scale, in the group enrolled in the MBI study compared to the group who refused to participate (q = 0.024).

**Table 2.**
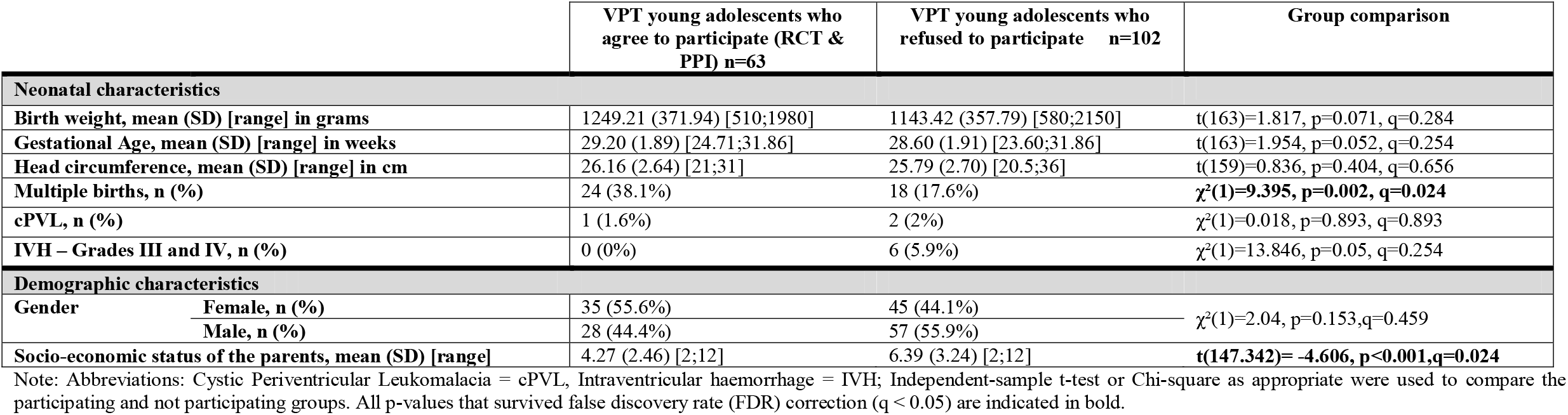
Demographic and neonatal characteristics of VPT young adolescents who were enrolled in the study (RCT and PPI designs together) and those who refused to participate

### Sample characteristics

Neonatal characteristics of participants enrolled in the study are shown in details in Supplementary Table S2. The mean birth weight was 1249.21 grams (SD = 371.94) ranging from 510 grams to 1980 grams, and the mean gestational age was 29.20 weeks (SD = 1.89) ranging from 24.71 to 31.86 weeks. The length of hospitalisation was on average of 61.87 days (SD = 29.68) ranging from 17 days to 151 days. Of the 63 participants, 24 were from multiple births (38.1%), 12 (19%) had bronchopulmonary dysplasia and no individuals suffered from intraventricular haemorrhage grade III or IV. Considering participants enrolled in the RCT only (n = 56), there were no significant differences between IG and WG for neonatal characteristics, including birth weight, gestational age, head circumference, length of hospitalisation as well as the presence of brain lesions and other medical conditions during the neonatal periods.

Demographic characteristics of participants enrolled in the study are shown in Supplementary Table S3. The mean age was 12.24 years (SD = 1.32) with 55.6% of females, and a mean GAI of 107.97 (SD = 11.15) ranging from 83 to 132. The mean Largo scale used to assess socio-economic status of the parents was 4.16 (SD = 2.43), where 2 is the highest possible socio-economic status and 12 the lowest. Parents’ nationality and native language are in accordance with the general population of the Geneva area with most parents being Swiss or French (71.4 %) and speaking French as their native language (about 88%). Considering participants enrolled in the RCT only (n = 56), there were no significant differences between IG and WG for the demographic characteristics, including gender, age, GAI, socio-economic status of the parents and parents’ nationalities and native language.

Self-reported as well as parents’ questionnaires were administered to gain further information on the participants’ medical history and global health, schooling and friendship (Supplementary Table S4 and Table S5). Overall, parents reported their young adolescents to be in good health (about 95% of all participants). Among neurodevelopmental diagnosis, 11.1% were diagnosed with Attention Deficit Hyperactivity Disorder (4.8% required psychostimulant treatment), 17.5% with dyslexia, dyspraxia or high intellectual capacity and 3.2% had motor disabilities. None of the participants had Autism Spectrum Disorder (ASD). Nevertheless, relatively high rates of participants needed, at some point, different types of interventions, including speech therapy (30.2%), physiotherapy (15.9%) and psychotherapy (20.6%). In terms of schooling, overall, most of the young adolescents felt competent in maths (66.7%), French literacy (66.7%) and sport (71.4%). Nevertheless, 33.3% of parents reported school difficulties and 27% of participants needed school support. About 78% of the participants reported typical social relationships, which corroborates parents’ assessment. Participants of our cohort were actively engaged in activities outside school including sport (58.7%) as well as art and music-related activities (44.4%).

### MBI satisfaction and attendance

The MBI sessions included 3 to 8 participants per group with an average of 5.2 participants per group. Thirty-seven VPT participants (71.2%) completed all the 8 sessions, 13 (25%) completed 7 sessions and 2 (3.8%) completed 6 sessions. None of the participants completed less than 6 out of the 8 MBI sessions. Based on the satisfactory questionnaire, participants rated the importance of this MBI program to them at 8.28 out of 10 (standard deviation = 1.8, range = 3 to 10)

## DISCUSSION

To our knowledge, this is the first study of MBI for prematurely born young adolescents using RCT and PPI designs. Taken together, our findings suggest that an MBI study I feasible to implement in VPT young adolescents and MBI is likely to be accepted and attractive to this vulnerable population and their families. Our study also provides detailed information on the MBI proposed by our group that was specifically developed to introduce mindfulness to young adolescents from 10 to 14 years of age.

Of young adolescents who were screened (n = 165), the final participation rate in the study was 38.2%, with 31.5% of participants who completed MBI and follow up assessments. Some families declined to participate due to lack of time, lack of interest, geographical constraints or unreachability. However, considering the high participant-load of this study, the final participation in the study can be considered as satisfactory. It is important to note that there was a significant difference in socio-economic status between the group who participated in the study and the one who refused (q = 0.024), with significantly higher socio-economic status in families who enrolled in the study. It is possible that families with higher socio-economic status have more resources to engage in such a commitment, considering the load induced by the study. Families with higher socio-economic status might also be more familiar with MBIs, which might increase the motivation and interest to participate in such a study. Therefore, due to the high socio-economic status of the families enrolled, our cohort might not be completely representative of the general VPT young adolescent population. It is possible that our cohort contains less vulnerable VPT young adolescents than the general population.

Compliance with study-related procedures was strong, including MBI sessions as well as assessments at the different time points. Once enrolled, only 17.5% of the participants dropped out before the start of the MBI due to lack of time, interest or unreachability. It highlights the importance of giving very detailed information to parents and families and to make sure that they have understood them properly. There was no drop-out once the participants started the MBI program. Of the participants who completed follow-up, we observed an average session attendance of 7.67 out of 8. Overall, the MBI was considered as important for the participants (mean of 8.28 out of 10). The majority of young adolescents took part in the RCT (92.3%) counting a total of four assessments (i.e., baseline, time 1, time 2, time 3), while a few joined the PPI design for time or geographical constraints (7.7%), counting a total of three assessments (i.e., baseline, pre-intervention assessment, post-intervention assessment). Participants enrolled in the study who followed the MBI completed all required assessments, including the MRI scan before and after MBI if there was no contra-indication. This shows that once enrolled in the study, and especially once the MBI started, participants and their families were interested and willing to commit to the intervention. Overall compliance results corroborate previous MBI studies completed in children and adolescents, showing a high rate of compliance once enrolled in the programme [28]. In regards to participants enrolled in the RCT, there was no significant difference overall between the IG and the WG for neonatal characteristics, including birth weight, gestational age, head circumference, length of hospitalisation, multiple birth and medical condition; as well as no significant difference for demographic characteristics, including age at baseline, socio-economic status and index of general ability.

Although satisfaction and attendance results are encouraging, this study should also be considered in the context of its limitation. Firstly, home mindfulness practice was not recorded in our study. We were therefore unable to have quantitative data on actual practice, an issue faced by many studies. For future research, it might be beneficial to collect precise quantitative data on their practice of mindfulness at home. For example, a phone application in which participants can access guided meditations and that is able to register data on individual usage might be a more reliable tool. Finally, although we found this intervention to be feasible, it may not be generalisable to other settings. For example, in the context of geographical constraints, families might not be able to commute for the MBI of their young adolescents. In addition, in our study, we were more likely to enrol young adolescents and families from higher socio-economic status, which could reflect that young adolescents from family with higher financial resources were more able to commit to such an intervention. Nevertheless, detailed sample characteristics observed in the current study (i.e., schooling, therapies) correspond to outcomes previously described in the preterm population [29].

## CONCLUSIONS

To our knowledge, this is the first study to investigate the feasibility of MBI study in prematurely born young adolescents. Our findings suggest that MBI study is feasible to implement. Once enrolled, the high rate of engagement in the intervention program also shows that participants largely accepted the MBI program. The use of an RCT design constitutes the gold standard for testing the efficacy of such intervention in VPT young adolescents and, if proven effective, MBI could potentially serve as a valuable tool for improving executive functions, behavioural and socio-emotional competences in this vulnerable population.

## Data Availability

Data are available on request to the PI: There are restrictions on data related to identifying participant information and appropriate ethical approval is required prior to release. Only de-identified data will be available.

## List of abbreviations

VPT: Very preterm
MBI: Mindfulness-based interventions
MRI: Magnetic Resonance Imaging
BPD: Neonatal bronchopulmonary dysplasia
IVH: Intraventricular haemorrhage
cPVL: Cystic periventricular leukomalacia
RCT: Randomised controlled trial
PPI: pre-post intervention
IG: intervention group
WG: waiting group
MBSR: Mindfulness-Based Stress Reduction
MBCT: Mindfulness-Based Cognitive Therapy
FDR: False Discovery Rate correction
WISC-IV: Wechsler Intelligence Scale for Children – 4th Edition
GAI: General Ability Index
MP-RAGE: Magnetization Prepared Rapid Gradient Echo
DWI: Diffusion-weighted imaging

## DECLARATIONS

### Ethics approval and consent to participate

This study was approved by the Swiss Ethics Committees on research involving humans (ID: 2015-00175). Written informed consent was obtained from from a parent or guardian and from the participant. The Magnetic Resonance Imaging (MRI) component to the study was optional and participants could still participate in the trial if the young adolescent or his/her family did not wish to have an MRI or if participants had a contra-indication to participate in the MRI session, such as dental braces.

### Consent for publication

Written informed consent has been obtained from the participant and the participant’s parent or guardian for the publication of this manuscript and any accompanying images.

### Competing interests

The authors declare that they have no competing interests.

### Funding

This study is funded by the Swiss National Science Foundation, No. 324730_163084 [PI: P.S. Hüppi]. The funding body did not have any role in this study’s design and conduct at any stage.

### Authors’ contributions

RHVL, CBT, AM, PSH, designed the study; DEM gave methodological advice; VS, MCL, LF, JDA, ES, FG collected the data; MMS, FSG, AM, RHVL, CBT performed the MBI interventions; VS, MCL wrote the paper; All authors reviewed and edited the paper.

## Acknowledgements

We thank and acknowledge all participating young adolescents and families who made this research possible. We also thank the Foundation Campus Biotech Geneva (FCBG), a foundation of the Swiss Federal Institute of Technology Lausanne (EPFL), the University of Geneva (UniGe), and the Hôpitaux Universitaires de Genève (HUG) for their practical help, in particular Roberto Martuzzi and Loan Mattera. We also thank the Plateforme de recherche pédiatrique of the HUG.

## ADDITIONAL FILES

**Supplementary Table S1**. Details of the research study’s neurobehavioural assessment

**Supplementary Table S2**. Neonatal characteristics of: (a) all young adolescents enrolled the study (RCT and PPI together), n=63); (b) young adolescents enrolled in the RCT only (n=56), as well IG and WG comparisons; (c) young adolescents enrolled in the PPI only (n=7).

**Supplementary Table S3**. Demographic characteristics at baseline of: (a) all young adolescents enrolled in the study (RCT and PPI together), n=63); (b) young adolescents enrolled in the RCT only (n=56), as well IG and WG comparisons; (c) young adolescents enrolled in the PPI only (n=7).

**Supplementary Table S4**. Parents’ questionnaire on their young adolescent schooling, friendship, medical history and global health

**Supplementary Table S5**. Self-reported questionnaire on schooling, friendship, activities and family organisation.

### Study Protocol

This file contains the study protocol.

### SPIRIT Checklist

In accordance with BioMed Central editorial policies, we completed the SPIRIT checklist as an additional file. This file contains the checklist in full with the page number/paragraph and section of your manuscript which reports the information that meets the criteria of the checklist.

### CONSORT Checklist

In accordance with BioMed Central editorial policies, we completed the CONSORT checklist as an additional file. This file contains the checklist in full with the page number/paragraph and section of your manuscript which reports the information that meets the criteria of the checklist.

## Notes

### Competing Interest Statement

The authors have declared no competing interest.

### Clinical Trial

NCT04638101

### Clinical Protocols

https://clinicaltrials.gov/show/NCT04638101

### Funding Statement

This study is funded by the Swiss National Science Foundation No. 324730_163084 [PI: P.S. Huppi].

### Author Declarations

This study was approved by the Swiss Ethics Committees on research involving humans (ID: 2015-00175). Written informed consent was obtained from the principal caregiver and from the participant. The Magnetic Resonance Imaging (MRI) component to the study was optional and participants could still participate in the trial if the young adolescent or his/her family did not wish to have an MRI or if participants had a contra-indication to participate in the MRI session, such as dental braces.

