## Supplementary Materials for "Improving executive, behavioural and socio-emotional competences in very preterm young adolescents through a mindfulness-based intervention: study protocol and feasibility"

**Supplementary Table S1.** Details of the research study's neurobehavioural assessment

| Domains | Modalities | Measures | Description |
|---|---|---|---|
| **Executive competences** | | | |
| | Parent questionnaire | | |
| | | Behaviour Rating Inventory of Executive Function, parent version (BRIEF) [1] | |
| | | | The BRIEF parent questionnaire provides an index of attention and executive abilities in everyday life. The BRIEF comprises 86 items over two index containing subscales: (i) Behavioural Regulation Index (BRI) comprising 3 subscores including, Inhibit, Shift, Emotional Control; (ii) Metacognition index (MCI) comprising 5 subscores including, Initiate, Working Memory, Plan/Organise, Organisation of Materials, Monitor; as well as a global score called the Global Executive Composite (GEC). |
| | Neuropsychological tests | | |
| | | Letter-Number Sequencing (WISC-IV) [2] | |
| | | | The letter-number sequencing is a working memory task. Sequences of number and letters are read to the participant, and he/she is then asked to re-sequence the numbers in numerical order from lowest to highest and then to sequence the letters in alphabetical order. |
| | | Tempo Test Rekenen [3] | |
| | | | The Tempo Test Rekenen is an arithmetic test consisting of 200 arithmetic number fact problems presented in five rows (one row with addition, one row with subtraction, one row with division, one row with multiplication, and one mixed problem row). Within each row, the problems increase in difficulty. Participant are asked to solve as many items as possible within 1 min per row. |
| | Neurocognitive computerised tasks | | |
| | | Flanker Visual Filtering Task [4] | |
| | | | The Flanker Visual Filtering Task was used to assess attentional control and information processing speed. Each trial showed a horizontal row of five fish. The participant was asked to respond as quickly as possible to whether the central fish was facing to the left or right. Congruent trials were the ones with all five fish in the horizontal row pointing in the same direction and incongruent trials were the ones with the four distracting fishes pointing in the opposite direction of the central target fish. |
| | | Reality Filtering Task [5] | |
| | | | The Reality Filtering task child-adapted version was used to assess recognition memory and orbitofrontal reality filtering. It consisted of a continuous recognition task composed of two runs with the same picture set but arranged in different order. The first run measures storage and recognition capacity (item memory), the second run measures reality filtering. |
| **Behaviour and socio-emotional competences** | | | |
| | Parent questionnaire | | |
| | | Strength and Difficulties Questionnaire, parent version (SDQ) [6] | |
| | | | The SDQ parent questionnaire assess overall behaviour problems, emotional symptoms, hyperactivity and inattention, peer relationship problems, and prosocial behaviour. It rates participant's behaviour over the previous 6 months. The SDQ is scored on a Likert scale and includes 25 items, providing five subscales: Emotional Symptoms, Conduct Symptoms, Hyperactivity-Inattention, Peer Problems, Prosocial Behaviour. A Total Difficulties score is also derived from the subscales. |
| | Self-reported questionnaires | | |

| | | |
|---|---|---|
| | KIDSCREEN-27 [7] | |
| | | The KIDSCREEN-27 is a self-reported questionnaire providing an index of health-related quality of life in children and adolescents. This instrument scored on a Likert scale and includes 27 items, providing five subscales: physical well-being (5 items), psychological well-being (7 items), autonomy and parent relation (7 items), social supports and peers (4 items), and school environment (4 items). |
| | Social Goal Scale (SGS)[8] | |
| | | The SGS is a self-reported questionnaire providing an index of social responsiveness and of goals setting which ultimately gets you involve with some social work. This instrument scored on a Likert scale and includes 11 items providing one score. |
| | Self-Compassion Scale – Short form (SCS) [9] | |
| | | The SCS is a self-reported questionnaire comprising 12 items, which produces a total global score that can also be classified into two subscores: negative behaviours toward the self (6 items) or positive behaviours towards the self (6 items). |
| Neuropsychological tests | | |
| | Affect Recognition (NEPSY-II) [10] | |
| | | The affect recognition subtest assesses the ability to recognise facial emotional expressions (happy, sad, anger, fear, disgust, and neutral) from photographs of children’s faces in several matching tasks. In the first task, the participant selected one of the four faces that depicted the same emotion as a child's face at the top of the page. In a second task, the participant selected two photographs of faces that displayed the same affect from a selection of four photographs. Finally, the participant examined a photograph of a child's face for 5 seconds, and then from memory, selected two photographs that matched the same emotion as the face previously shown. |
| | Theory of Mind (NEPSY-II) [10] | |
| | | The theory of mind subtest measures understanding of mental functions and other people's perspectives.<br>In the first task, questions are asked to the participant about different verbal scenarios measuring understanding of beliefs, intentions, others’ thoughts, ideas and comprehension of figurative language. In the second task, participants have to match facial emotional expressions, from photographs of children’s faces, to a scenario. |
| Neurocognitive computerised tasks | | |
| | Emotion Regulation task [11] | |
| | | The Emotion Regulation task was used to assessed emotional regulation capacities. Participants were asked to watch positive, negative and neutral film clips, after which they were asked to calm down and concentrate on their breath. They had to rate the film clips from 1 (positive) to 5 (negative) on a Likert scale. Film clips videotaped by amateurs were taken from Samson and colleagues (2016).[11] During this task, physiological responses (heart rate, skin conductance and respiration) were acquired in the MRI scanner as well as in the simulated “mock” MRI. |
| | Recognition of Emotion in Contextual Scene Task [12] | |
| | | The Recognition of Emotion in Contextual Scene task was used to assessed facial emotional expression recognition based on contextual and facial cues. Participants were first presented with a visual scene from one of the two condition: (a) scene that included a character interacting with a peer or with meaningful objects, called context condition; and (b) scene that included a character but in the absence of any peers or meaningful objects, called no-context condition. Three emotional facial expressions among anger, disgust, happiness, fear, and sadness were then presented to the participant who had to choose the emotion felt by the character in the scene. |

**Supplementary Table S2.** Neonatal characteristics of: (a) all young adolescents enrolled the study (RCT and PPI together), n=63); (b) young adolescents enrolled in the RCT only (n=56), as well IG and WG comparisons; (c) young adolescents enrolled in the PPI only (n=7).

| | All young adolescents enrolled (RCT and PPI), n=63 | RCT, n=56 | | | PPI, n=7 |
|---|---|---|---|---|---|
| | | Intervention group (IG) n=29 | Waiting group (WG) n=27 | Group comparison (IG vs WG) | |
| **Birth weight, mean (SD) [range] in grams** | 1249.21 (371.94) [510;1980] | 1284.83 (351.41) [650;1810] | 1210 (400.85) [520;1980] | t(54)=0.744, p=0.460, q= 0.656 | 1252.86 (379.24) [510;1680] |
| **Gestational age, mean (SD) [range] in weeks** | 29.20 (1.89) [24.71;31.86] | 29.29 (1.92) [24.71; 31.86] | 29.12 (1.93) [26;31.71] | t(54)=0.317 p=0.753, q= 0.795 | 29.04 (2.01) [25.71;30.86] |
| **Head circumference, mean (SD) [range] in cm** | 26.16 (2.64) [21;31] | 26.55 (2.57) [21;31] | 25.65 (2.82) [21;31] | t(53)=1.234, p=0.223, q= 0.595 | 26.46 (2.21) [22;29] |
| **Length of hospitalisation, mean (SD) [range] in days** | 61.87 (29.68) [17;151] | 59.56 (26.79) [23;131] | 63 (33.69) [17;151] | t(52)=-0.416,p=0.679, q= 0.776 | 66.43 (26.96) [36;123] |
| **Multiple births, n (%)** | 24 (38.1%) | 13 (44.8%) | 7 (25.9%) | $\chi^2(2)$=2.202, p=0.333, q= 0.656 | 4 (57.1%) |
| **cPVL, n(%)** | 1 (1.6%) | 1 (3.4%) | 0 | $\chi^2(1)$=0.903, p=0.342, q= 0.656 | 0 |
| **IVH - Grades III and IV, n (%)** | 0 (0%) | 0 (0%) | 0 (0%) | - | 0 |
| **BPD, n (%)** | 12 (19%) | 5 (17.2%) | 6 (22.2%) | $\chi^2(1)$=0.534, p=0.465, q=0.656 | 1 (14.3%) |

Note: Abbreviations: Cystic Periventricular Leukomalacia = cPVL, Intraventricular haemorrhage = IVH, Bronchopulmonary dysplasia = BPD; Independent-sample t-test, Chi-square as appropriate were used to compare the randomised groups. All p-values that survived false discovery rate (FDR) correction (q < 0.05) are indicated in bold.

**Supplementary Table S3.** Demographic characteristics at baseline of: (a) all young adolescents enrolled in the study (RCT and PPI together), n=63); (b) young adolescents enrolled in the RCT only (n=56), as well IG and WG comparisons; (c) young adolescents enrolled in the PPI only (n=7).

| | | All young adolescents enrolled (RCT and PPI), n=63 | RCT, n=56 | | | PPI, n=7 |
|---|---|---|---|---|---|---|
| | | | Intervention group (IG), n=29* | Waiting group (WG), n=27* | Group comparison (IG vs WG) | |
| **Gender** | **Female, n** | 35 (55.6%) | 14 (48.3%) | 16 (59.3%) | $\chi^2(1)=0.678$, p=0.410, q= 0.656 | 5 (71.4%) |
| | **Male, n** | 28 (44.4%) | 15 (51.7%) | 11 (40.7%) | | 2 (28.6%) |
| **Age at baseline, mean (SD) [range] in years** | | 12.24 (1.32) [10.08;14.85] | 12.05 (1.23) [10.08;14.24] | 12.26 (1.37) [10.38;14.85] | t(50)=-0.585, p=0.561, q= 0.692 | 13 (1.49) [11.59;14,84] |
| **Index of general ability (GAI), mean (SD) [range]** | | 107.97 (11.15) [83;132] | 106.67 (11.47) [83;132] | 108.76 (11.23) [87;130] | t(50)=-0.664, p=.0.510, q=0.680 | 110.5 (10.31) [101;127] |
| **Socio-economic status (SES), mean (SD) [range]** | | 4.16 (2.43) [2;12] | 4.78 (2.62) [2;12] | 3.76 (2.35) [2;12] | t(50)=1.470, p=0.148, q= 0.459 | 3 (0.894) [2;4] |
| **Mother's nationality** | **Swiss, n (%)** | 30 (47.6%) | 17 (58.6%) | 12 (44.4%) | $\chi^2(2)=1.606$, p=0.448, q= 0.656 | 1 (14.3%) |
| | **French, n (%)** | 15 (23.8%) | 6 (20.7%) | 6 (22.2%) | | 3 (42.9%) |
| | **Other, n (%)** | 13 (22.4%) | 4 (13.8%) | 7 (25.9%) | | 2 (28.6%) |
| | **Unknown, n (%)** | 5 (7.9%) | 2 (6.9%) | 2 (7.4%) | | 1 (14.3%) |
| **Father's nationality** | **Swiss, n (%)** | 29 (46%) | 14 (48.3%) | 12 (44.4%) | $\chi^2(2)=0.545$, p=0.762, q= 0.795 | 3 (42.9%) |
| | **French, n (%)** | 16 (25.4%) | 7 (24.1%) | 6 (22.2%) | | 3 (42.9%) |
| | **Other, n (%)** | 12 (19%) | 5 (17.2%) | 7 (25.9%) | | 0 |
| | **Unknown, n (%)** | 5 (7.9%) | 3 (10.3%) | 2 (7.4%) | | 1 (14.3%) |
| **Mother's native language** | **French, n (%)** | 38 (60.3%) | 18 (62.1%) | 17 (63%) | $\chi^2(1)=0.311$, p=0.577, q= 0.692 | 3 (42.9%) |
| | **Other, n (%)** | 18 (28.6%) | 9 (31%) | 6 (22.2%) | | 3 (42.9%) |
| | **Unknown, n (%)** | 7 (11.1%) | 2 (6.9%) | 4 (14.8%) | | 1 (14.3%) |
| **Father's native language** | **French, n (%)** | 40 (63.5%) | 19 (65.5%) | 15 (55.6%) | $\chi^2(1)=1.051$, p=0.305, q= 0.656 | 6 (85.7%) |
| | **Other, n (%)** | 15 (23.8%) | 6 (20.7%) | 9 (33.3%) | | 0 |
| | **Unknown, n (%)** | 8 (12.7%) | 4 (13.8%) | 3 (11.1%) | | 1 (14.3%) |

Note: *there was missing data for the following measures: (i) age at baseline, IG n=27, WG n=25, PPI n=6; (ii) Index of general ability, IG n=27, WG n=25, PPI n=6; (iii) SES, IG n=27, WG n=25, PPI n=6. Independent-sample t-test or Chi-Square were used to compare the intervention and the waiting randomised groups. All p-values that survived false discovery rate (FDR) correction (q < 0.05) are indicated in bold.

**Supplementary Table S4.** Parents' questionnaire on their young adolescent schooling, friendship, medical history and global health

| | | All young adolescents enrolled (RCT and PPI), n=63 | RCT, n=56 | | PPI, n=4 |
|---|---|---|---|---|---|
| | | | Intervention group (IG), n=29 | Waiting group (WG), n=27 | |
| **Repeated a school year** | **yes** | 6 (9.5%) | 3 (10.3%) | 2 (7.4%) | 1 (14.3%) |
| | **unknown** | 7 (11.1%) | 2 (6.9%) | 4 (14.8%) | 1 (14.3%) |
| **Skipped a grade** | **yes** | 2 (3.2%) | 1 (3.4%) | 0 (0.0%) | 1 (14.3%) |
| | **unknown** | 7 (11.1%) | 2 (6.9%) | 4 (14.8%) | 1 (14.3%) |
| **Friends' number** | **2 or less** | 9 (14.3%) | 5 (17.2%) | 4 (14.8%) | 0 (0.0%) |
| | **more than 2** | 45 (71.4%) | 21 (72.4%) | 19 (70.4%) | 5 (71.4%) |
| | **unknown** | 9 (14.3%) | 3 (10.3%) | 4 (14.8%) | 2 (28.6%) |
| **Motor disabilities** | **yes** | 2 (3.2%) | 1 (3.4%) | 1 (3.7%) | 0 |
| | **unknown** | 7 (11.1%) | 2 (6.9%) | 4 (14.8%) | 1 (14.3%) |
| **Attention Deficit Hyperactivity Disorder** | **yes** | 7 (11.1%) | 4 (13.8%) | 2 (7.4%) | 1 (14.3%) |
| | **unknown** | 7 (11.1%) | 2 (6.9%) | 4 (14.8%) | 1 (14.3%) |
| **School difficulties** | **yes** | 21 (33.3%) | 9 (31%) | 10 (37%) | 2 (28.6%) |
| | **unknown** | 9 (14.3%) | 2 (6.9%) | 5 (18.5%) | 2 (28.6%) |
| **School support** | **yes** | 17 (27%) | 7 (24.1%) | 10 (37%) | 0 |
| | **unknown** | 8 (12.7%) | 2 (6.9%) | 4 (14.8%) | 2 (28.6%) |
| **Neurodevelopmental diagnosis** (options include dyslexia, dysorthographia, dyscalculia, dysgraphia, dyspraxia, high intellectual capacities, autism spectrum disorder) | **reported** | 11 (17.5%) | 6 (20.7%; dyslexia n=2, dyspraxia, n=1, high intellectual capacities n=1, unknown n=2) | 3 (11.1%; dyslexia n=2, high intellectual capacities n=1) | 2 (28.6%; dyslexia n=1, unknown n=1) |
| | **unknown** | 8 (12.7%) | 2 (6.9%) | 4 (14.8%) | 2 (28.6%) |
| **Interventions** | **speech therapy** | 19 (30.2%) | 7 (24.1%) | 10 (37%) | 2 (28.6%) |
| | **speech therapy ongoing** | 5 (7.9%) | 0 (0.0%) | 3 (11.1%) | 2 (28.6%) |
| | **physiotherapy** | 10 (15.9%) | 3 (10.3%) | 6 (22.2%) | 1 (14.3%) |
| | **physiotherapy ongoing** | 1 (1.6%) | 0 (0.0%) | 1 (3.7%) | 0 |
| | **psychomotor therapy** | 8 (12.7%) | 3 (10.3%) | 3 (11.1%) | 2 (28.6%) |
| | **psychomotor therapy ongoing** | 2 (3.2%) | 0 (0.0%) | 0 (0.0%) | 2 (28.6%) |
| | **occupational therapy** | 7 (11.1%) | 2 (6.9%) | 4 (14.8%) | 1 (14.3%) |

| | | | | | |
|---|---|---|---|---|---|
| | **occupational therapy ongoing** | 3 (4.8%) | 2 (6.9%) | 0 (0.0%) | 1 (14.3%) |
| | **psychotherapy** | 13 (20.6%) | 8 (27.6%) | 4 (14.8%) | 1 (14.3%) |
| | **psychotherapy ongoing** | 5 (7.9%) | 3 (10.3%) | 1 (3.7%) | 1 (14.3%) |
| | **unknown** | 8 (12.7%) | 2 (6.9%) | 4 (14.8%) | 2 (28.6%) |
| **Ongoing medication** | **psychostimulant treatment** | 3 (4.8%) | 2 (6.9%) | 0 (0.0%) | 1 (14.3%) |
| | **growth hormones** | 1 (1.6%) | 0 | 1 (4.5%) | 0 |
| | **unknown** | 7 (11.1%) | 2 (6.9%) | 4 (14.8%) | 1 (14.3%) |
| **Health** | **participant not in good health** | 3 (4.8%) | 1 (3.4%) | 2 (7.4%) | 0 |
| | **participant wears glasses** | 25 (39.7%) | 12 (41.4%) | 10 (37%) | 3 (42.9%) |
| | **hearing aid** | 1 (1.6%) | 0 (0.0%) | 1 (3.7%) | 0 (0.0%) |
| | **unknown** | 8 (12.7%) | 2 (6.9%) | 4 (14.8%) | 2 (28.6%) |
| **Participant already took part in Mindfulness program** | | 0 | 0 | 0 | 0 |

Note: If psychostimulant treatment was an ongoing medication, the treatment was taken by the participant consistently throughout the study and during the assessments.

**Supplementary Table S5.** Self-reported questionnaire on schooling, friendship, activities and family organisation

| | | All young adolescents enrolled (RCT and PPI), n=63 | RCT, n=56 | | PPI, n=7 |
|---|---|---|---|---|---|
| | | | Intervention group (IG), n=29 | Waiting group (WG), n=27 | |
| **School degree (Swiss system)** | **$6^{th}$ grade** | 2 (3.2%) | 0 (0%) | 1 (3.7%) | 1 (14.3%) |
| | **$7^{th}$ grade** | 12 (19%) | 6 (20.7%) | 6 (22.2%) | 0 (0%) |
| | **$8^{th}$ grade** | 19 (30.2%) | 11 (37.9%) | 7 (25.9%) | 1 (14.3%) |
| | **$9^{th}$ grade** | 13 (20.6%) | 4 (13.8%) | 6 (22.2 %) | 3 (42.9%) |
| | **$10^{th}$ grade** | 4 (6.3%) | 3 (10.3%) | 1 (3.7%) | 0 (0%) |
| | **$11^{th}$ grade** | 6 (9.5%) | 2 (6.9%) | 3 (11.1%) | 1 (14.3%) |
| | **unknown** | 7 (11.1%) | 3 (10.3%) | 3 (11.1%) | 1 (14.3%) |
| **Feel competent in maths?** | **yes** | 42 (66.7%) | 21 (72.4%) | 17 (63%) | 4 (57.1%) |
| | **no** | 14 (22.2%) | 5 (17.2%) | 7 (25.9%) | 2 (28.6%) |
| | **unknown** | 7 (11.1% | 3 (10.3%) | 3 (11.1%) | 1 (14.3%) |
| **Feel competent in French literacy?** | **yes** | 42 (66.7%) | 19 (65.5%) | 19 (70.4%) | 4 (57.1%) |
| | **no** | 14 (22.2%) | 7 (24.1) | 5 (18.5%) | 2 (28.6%) |
| | **unknown** | 7 (11.1%) | 3 (10.3%) | 3 (11.1%) | 1 (14.3%) |
| **Feel competent in sport?** | **yes** | 45 (71.4%) | 19 (65.5%) | 20 (74.1%) | 6 (85.7%) |
| | **no** | 11 (17.5%) | 7 (24.1%) | 4 (14.8%) | 0 (0%) |
| | **unknown** | 7 (11.1%) | 3 (10.3%) | 3 (11.1%) | 1 (14.3%) |
| **Do you have a best friend?** | **yes** | 49 (77.8%) | 23 (79.3%) | 20 (74.1%) | 6 (85.7%) |
| | **no** | 6 (9.5%) | 3 (10.3%) | 3 (11.1%) | 0 (0%) |
| | **unknown** | 8 (12.7%) | 3 (10.3%) | 4 (14.8%) | 1 (14.3%) |
| **Activities outside school** | **sport** | 37 (58.7%) | 18 (62.1%) | 15 (55.6%) | 4 (57.1%) |
| | **art & music** | 28 (44.4%) | 12 (48.3%) | 10 (37%) | 6 (85.7%) |
| | **other** | 10 (15.9%) | 3 (10.3%) | 7 (25.9%) | 0 (0%) |
| | **unknown** | 7 (11.1%) | 3 (10.3%) | 3 (11.1%) | 1 (14.3%) |
| **Participant living with:** | **2 biological parents** | 45 (71.4%) | 20 (69%) | 19 (70.4%) | 6 (85.7%) |
| | **1 biological parent** | 10 (15.9%) | 5 (17.2%) | 5 (18.5%) | 0 (0%) |
| | **other** | 1 (1.6%) | 1 (3.4%) | 0 (0%) | 0 (0%) |
| | **unknown** | 7 (11.1%) | 3 (10.3%) | 3 (11.1%) | 1 (14.3%) |
| **Number of siblings** | **single child** | 3 (4.8%) | 1 (3.4%) | 1 (3.7%) | 1 (14.3%) |
| | **1 sibling** | 28 (44.4%) | 13 (44.8%) | 14 (51.9%) | 1 (14.3%) |
| | **2 siblings** | 20 (31.7%) | 9 (31%) | 6 (22.2%) | 5 (71.4%) |

| | | | | | |
|---|---|---|---|---|---|
| | **3 siblings** | 4 (6.3%) | 1 (3.4%) | 3 (11.1%) | 0 (0%) |
| | **4 siblings** | 1 (1.6%) | 1 (3.4%) | 0 (0%) | 0 (0%) |
| | **Unknown** | 7 (11.1%) | 4 (13.8%) | 3 (11.1%) | 0 (0%) |
| **Language spoken at home** | **French** | 49 (77.8%) | 24 (82.8%) | 19 (70.4%) | 6 (85.7%) |
| | **other** | 7 (11.1%) | 2 (6.9%) | 5 (18.5%) | 0 (0%) |
| | **unknown** | 7 (11.1%) | 3 (10.3%) | 3 (11.1%) | 1 (14.3%) |
| **Preferred spoken language** | **French** | 47 (74.6%) | 24 (82.8%) | 17 (63%) | 6 (85.7%) |
| | **other** | 9 (14.3%) | 2 (6.9%) | 7 (25.9%) | 0 (0%) |
| | **unknown** | 7 (11.1%) | 3 (10.3%) | 3 (11.1%) | 1 (14.3%) |
